# The QT Interval in Patients with SARS-CoV-2 Infection Treated with Hydroxychloroquine/Azithromycin

**DOI:** 10.1101/2020.04.02.20047050

**Authors:** Ehud Chorin, Matthew Dai, Eric Shulman, Lalit Wadhwani, Roi-Bar-Cohen, Chirag Barbhaiya, Anthony Aizer, Douglas Holmes, Scott Bernstein, Michael Spinelli, David S. Park, Larry A. Chinitz, Lior Jankelson

**Author notes:** **Corresponding authors:** Lior Jankelson MD PhD and Ehud Chorin MD PhD. **Conflicts of interest:** None. **Financial Disclosures:** None.

## Abstract

We report the change in the QT interval in 84 adult patients with SARS-CoV-2 infection treated with Hydroxychloroquine/Azithromycin combination. QTc prolonged maximally from baseline between days 3 and 4. in 30% of patients QTc increased by greater than 40ms. In 11% of patients QTc increased to >500 ms, representing high risk group for arrhythmia. The development of acute renal failure but not baseline QTc was a strong predictor of extreme QTc prolongation.

## Main

The SARS-CoV-2 pandemic is resulting in a staggering worldwide morbidity and mortality currently estimated at more than 650,000 positive cases and more than 30,000 confirmed deaths as of March 28, 2020 [1]. Although there are no approved drugs to prevent or treat SARS-CoV-2 infection, a recent report suggested that the combination of Hydroxychloroquine with Azithromycin (HY/AZ) may have favorable effect on clinical outcomes and viral load of infected patients [2]. This, has resulted in massive adoption of the regimen by clinicians worldwide. However, both medications have been independently shown to increase the risk for QT interval prolongation, drug-induced torsades de pointes (TdP), and drug induced-sudden cardiac death (SCD) [3-6]. There is no data regarding the effect of HY/AZ on the QT interval and the risk for malignant arrhythmia induction. Given the vast adoption of the regimen, including for post exposure prophylaxis [7], and the inherent risk for cardiac involvement in SARS-CoV-2 disease [8], we report here the change in QTc interval and the risk for TdP in 84 consecutive patients admitted to our institution with SARS-CoV-2 infection and treated with HY/AZ.

The clinical and epidemiological characteristics are presented in Table 1. Four patients died from multi-organ failure, without evidence of arrhythmia. There were no TdP events recorded, including in patients with severely prolonged QTc, defined as greater than 500 ms. The average time of ECG follow up post HY/AZ exposure was 4.3 +/- 1.7 days. The QTc interval prolonged from a baselines of 435 ± 24 ms to a maximal value of 463 ± 32 ms (p<0.001) which occurred on day 3.6 ± 1.6 of therapy. The change in daily QTc interval is presented in Figure 1. Of note, 11% of patients developed new severe QTc prolongation of > 500ms. On multivariate analysis, the development of acute renal failure was a significant predictor of severe QTc prolongation. Baseline Creatinine and Coadministration of Amiodarone trended for significance, as opposed to other QT prolonging medications. Importantly, baseline QTc and QTc > 460 ms did not predict QTc prolongation (supplementary table 1). The effectiveness of Hydroxychloroquine in treating SARS-CoV-2 infection was demonstrated in several invitro studies [9, 10] and in one small human study [2, 11]. However, given the lack of effective therapy, this combination is used liberally across the world and is recommended in multiple guidelines [12]. In this preliminary work we found that the in patients treated with HY/AZ QTc prolonged significantly. In 11% of patients, QTc prolonged to >500 ms, a known marker for high risk of malignant arrhythmia and sudden cardiac death [13-15]. Of note, recent guidance suggested screening of candidate patients for novel SARS-CoV-2 therapies, including HY/AZ based on baseline QTc [16]. Our data suggest that baseline QTc is not a reliable predictor of severe QTc prolongation in these patients. We suggest that QTc should be followed repeatedly in patients with SARS-CoV-2 infection treated with HY/AZ, particularly in patients with renal failure, a common complication in patients with SARS-CoV-2.

**Table 1:** Baseline clincial and epidemiological characteristics (n=84)

|  |  |
| --- | --- |
| Age | 63 ± 15 |
| Gender (% male) | 74% |
| Weight (Kg) | 86 ± 23 |
| Coronary artery disease | 11% |
| Hypertension | 65% |
| Chronic kidney disease | 7% |
| Diabetes mellitus | 20% |
| Chronic obstructive pulmonary disease | 8% |
| Congestive heart failure | 2% |
| Creatinine at initiation (mg%) | 1.4 ± 1.4 |
| Creatinine at max QTc (mg%) | 1.5 ± 1.4 |
| Acute renal failure | 6% |
| Abnormal LFTs at initiation | 33% |
| Abnormal LFTs at max QTc | 40% |
| SSRI/SNRI, lithium, or Lacosamide | 11% |
| Levofloxacin, Lopinavir/Ritonavir, or Tacrolimus | 8% |
| Norepinephrine, Phenylephrine, or Vasopressin | 13% |
| Amiodarone | 7% |
| Baseline QTc (ms) | 435 ± 24 |
| Maximum QTc (ms) | 463 ± 32 |
| Day of maximum QTc | 3.6 ± 1.6 |

**Figure 1:**
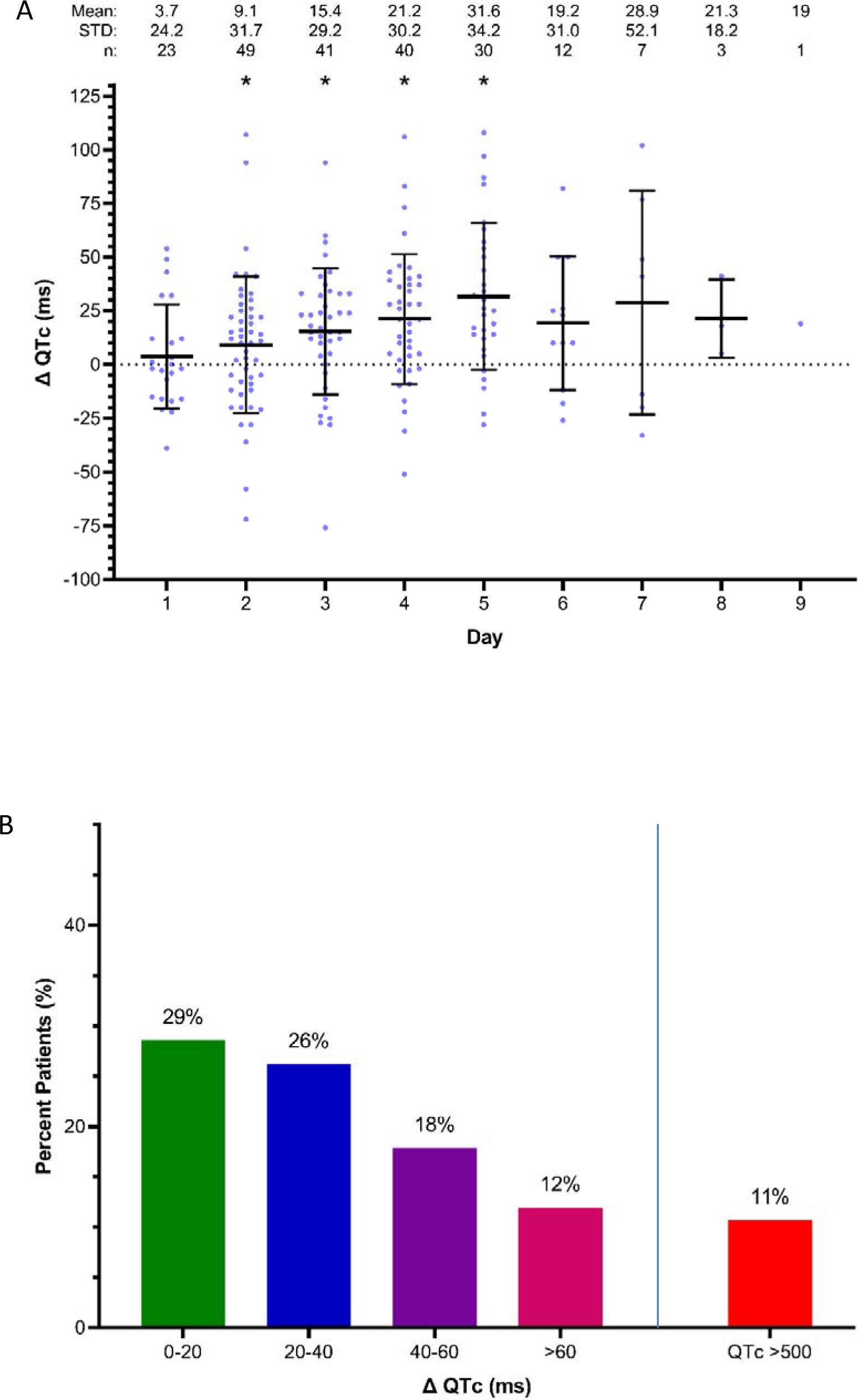
A, the change in QTc in days post HY/AZ initiation. *p <0.01 for QTc as compared to baseline QTc. B. percent patients with ranges of QTc prolongation.

**Supplementary table 1.** Predictors of maximal QTc >500.

| Univariate Logistic Regressions |  |  |  |
| --- | --- | --- | --- |
| Variable | *p* value | OR | 95% CI |
| Age | 0.43 | 0.98 | 0.94-1.03 |
| Gender (% male) | 0.30 | 0.32 | 0.04-2.73 |
| Weight | 0.72 | 1.01 | 0.98-1.04 |
| Coronary artery disease | 0.25 | 2.78 | 0.48-16.03 |
| Hypertension | 0.42 | 1.97 | 0.38-10.16 |
| Chronic kidney disease | 0.09 | 5.07 | 0.78-32.79 |
| Diabetes mellitus | 0.31 | 2.18 | 0.49-9.79 |
| Chronic obstructive pulmonary disease | 1 | - | - |
| Congestive heart failure | 0.13 | 9.25 | 0.53-162.52 |
| Creatinine at initiation | <0.01 | 1.76 | 1.15-2.7 |
| Creatinine at max QTc | <0.01 | 1.75 | 1.2-2.54 |
| Acute renal failure | <0.01 | 18.25 | 2.54-131.3 |
| Abnormal LFTs at initiation | 0.29 | 0.31 | 0.04-2.75 |
| Abnormal LFTs at max QTc | 0.12 | 0.18 | 0.02-1.59 |
| SSRI/SNRI, Lithium, or lacosamide | 0.97 | 1.05 | 0.12-9.49 |
| Levofloxacin, lopinavir/ritonavir, or tacrolimus | 1 | - | - |
| Amiodarone | <0.01 | 12.00 | 1.98-72.89 |
| Baseline QTc | 0.10 | 1.03 | 1.00-1.06 |
| Baseline QTc > 460 ms | 0.03 | 5.20 | 1.19 - 22.7 |
| Multivariate Logistic Regression |  |  |  |
| Variable | *p* value | OR | 95% CI |
| Creatinine at initiation (mg/dL) | 0.07 | 1.52 | 0.97 - 2.37 |
| Acute renal failure | 0.01 | 19.45 | 2.06 - 183.38 |
| Amiodarone | 0.09 | 7.06 | 0.70 - 71.79 |
| Baseline QTc > 460 ms | 0.28 | 2.90 | 0.42 - 20.24 |

## Methods

This is a retrospective study performed at NYU Langone medical center, New York. We reviewed 84 consecutive adult patients who were hospitalized at NYU Langone medical center with a positive SARS-CoV-2 disease and were treated with the combination of HY/AZ. Medical records were reviewed to obtain epidemiologic characteristics, comorbidities, baseline off-drug ECGs as well as on-drugs ECGs. The closing date of follow-up was March 28th 2020. The study was performed according to our Institutional Review Board guidance in accordance with the ethical standards laid down in the 1964 Declaration of Helsinki and its later amendments, with a waiver of informed consent. The study was approved by the IRB committee of NYU Langone Health and NYU School of Medicine.

For data collection, Microsoft Excel (MS Excel 2013, v.15.0) was used for collection of the epidemiological and clinical information.

Statistical analysis was performed using IBM SPSS Statistics 26, and figures were constructed using GraphPad Prism 8. Continuous variables are expressed as mean ± standard deviation, and categorical variables are expressed as percentages. Normality of data samples was assessed using Shapiro-Wilk test and visual analysis of Q-Q plots. Hypothesis testing for comparing baseline QTc and maximum QTc was performed using paired samples t-test. For Figure 1, one sample t-test was performed to compare each sample against a delta QTc of 0ms (ie. no change from baseline). For Table 2, univariate and multivariate logistic regression was performed to identify predictors of maximum QTc >500ms. For Figure 3, a receiver operating characteristics (ROC) curve was calculated to evaluate the predictive value of baseline QTc for maximum QTc >500ms.

## Data Availability

We will make data available after approval of data committee

## Data availability

The data in this study will be shared upon request and approval will be designated by a data access committee. The data access committee comprises four corresponding authors and there is no restriction to data access.

## Contributions

L.J and E.C contributed to the study design and data interpretation and writing of manuscript. R.B, E.S and L.W contributed to data collection analysis. M.D contributed to statistical analysis. A.A, M.S, D.H, D.P, S.B and L.C contributed to critical revision of the manuscript. All authors reviewed and approved the final version of the manuscript.

## Notes

### Competing Interest Statement

The authors have declared no competing interest.

### Funding Statement

No external funding provided

